# Increasing Sample Diversity in Psychiatric Genetics – Introducing a new Cohort of Patients with Schizophrenia and Controls from Vietnam – Results from a Pilot Study

**DOI:** 10.1101/2021.04.21.21255615

**Authors:** VT Nguyen, A Braun, J Kraft, TMT Ta, GM Panagiotaropoulou, VP Nguyen, TH Nguyen, V Trubetskoy, CT Le, TTH Le, XT Pham, I Heuser-Collier, NH Lam, K Böge, IM Hahne, M Bajbouj, MM Zierhut, E Hahn, S Ripke

## Abstract

**Objectives:** Genome-Wide Association Studies (GWAS) of Schizophrenia (SCZ) have provided new biological insights; however, most cohorts are of European ancestry. As a result, derived polygenic risk scores (PRS) show decreased predictive power when applied to populations of different ancestries. We aimed to assess the feasibility of a large-scale data collection in Hanoi, Vietnam, contribute to international efforts to diversify ancestry in SCZ genetic research and examine the transferability of SCZ-PRS to individuals of Vietnamese Kinh ancestry.

**Methods:** In a pilot study, 368 individuals (including 190 SCZ cases) were recruited at the Hanoi Medical University’s associated psychiatric hospitals and outpatient facilities. Data collection included sociodemographic data, baseline clinical data, clinical interviews assessing symptom severity and genome-wide SNP genotyping. SCZ-PRS were generated using different training data sets: i) European, ii) East-Asian and iii) trans-ancestry GWAS summary statistics from the latest SCZ GWAS meta-analysis.

**Results:** SCZ-PRS significantly predicted case status in Vietnamese individuals using mixed-ancestry (R^2^ liability=4.9%, p=6.83*10^−8^), East-Asian (R^2^ liability=4.5%, p=2.73*10^−7^) and European (R^2^ liability=3.8%, p = 1.79*10^−6^) discovery samples.

**Discussion:** Our results corroborate previous findings of reduced PRS predictive power across populations, highlighting the importance of ancestral diversity in GWA studies.

## 1 Introduction

Schizophrenia (SCZ) is a severe and often chronic psychiatric disorder that consistently affects between 0,5-1% of the worldwide population (McGrath et al., 2008). It is a clinically heterogeneous disorder characterised by perceptual distortions such as delusions and hallucinations, disorganised thinking, negative symptoms including reduced affective resonance, avolition, lack of motivation, marked cognitive deficits, as well as mood disturbances (Galderisi et al., 2018). With a high disease burden (GBD 2016 Disease and Injury Incidence and Prevalence Collaborators, 2017), loss of social functioning, and reduced life-expectancy (Walker et al., 2015), SCZ has a substantial impact on public health-related costs (Cloutier et al., 2016). To this date, SCZ remains challenging to treat, as only about 13.5% of patients meet the criteria for full recovery (Charlson et al., 2018), and advances in treatment options have been hampered — at least in part — by our incomplete knowledge of SCZ etiology and pathophysiological mechanisms.

As with all complex traits and diseases, schizophrenia exhibits a diffuse pattern of underlying genetic variation in which many common genetic variants contribute to disease risk. In recent years large-scale genome-wide association studies (GWAS) have provided robust evidence for many susceptibility loci within the genome (Schizophrenia Working Group of the Psychiatric Genomics Consortium, 2014); the most recent SCZ GWAS meta-analysis identified 276 distinct genomic regions (Schizophrenia Working Group of the Psychiatric Genomics Consortium et al., 2020). While genetic studies can provide insights into the biological underpinnings of SCZ, they also offer potential biomarkers that could inform treatment strategies, patient management, prognosis, and possibly early intervention. A growing number of studies have directed their attention towards the utility of polygenic risk scores (PRS) in clinical psychiatry. PRS are typically constructed using genome-wide summary statistics and represent an individual’s propensity towards certain traits or disorders attributable to common genetic variation (International Schizophrenia Consortium et al., 2009). To establish the clinical relevance of genetic findings, it becomes increasingly important to invest in large, well-phenotyped SCZ cohorts; especially as patients affected by severe psychotic disorders are often underrepresented in population-based research initiatives (J. Martin et al., 2016)

Although increases in sample size boost statistical power, and consequently the discovery of novel risk-conferring variants (Levinson et al., 2014) and predictive capacity of PRS, cohorts of non-European ancestries — especially those from low- and middle-income countries — are still sparse in GWA studies. Multiple works have demonstrated that the prediction accuracy of PRS depends on the ancestral composition of discovery cohorts and their genetic divergence from the target sample (Bigdeli et al., 2020; Lam et al., 2019). As a result, polygenic scores derived from Eurocentric GWAS suffer from an attenuated predictive power when transferred to non-European populations (Duncan et al., 2019; A. R. Martin et al., 2019). Consequently, PRS are less informative for non-European populations; hence, patients from these populations will benefit less from polygenic scores and their potential applications. Although there is emerging evidence for a shared genetic architecture of schizophrenia across diverse populations (Lam et al., 2019), the incomplete transferability of GWAS findings is driven by three fundamental factors: 1) differences in allele frequencies across populations, as GWAS are powered only for common variants of small or modest effect-size; 2) differences in linkage disequilibrium patterns that affect how well causal variants are tagged by genotype arrays and imputation reference panels; 3) population genetic factors such as genetic drift, polygenic selection, and demographic history (Duncan et al., 2019; A. R. Martin et al., 2017, 2019). To improve GWAS findings’ generalizability, encouraging ancestral diversity in study samples has taken high priority in the field of psychiatric genetics (Mills & Rahal, 2020).

Despite a surge of East-Asian samples in the latest schizophrenia GWAS (Schizophrenia Working Group of the Psychiatric Genomics Consortium et al., 2020), a representative Vietnamese cohort is still lacking. To close this gap, we conducted a case-control pilot study in Hanoi, Vietnam, in which sociodemographic-, genetic-, and clinical data from a total of 378 participants were collected. Here we provide a first report on data collection’s feasibility while providing insights into clinical data and current treatment of patients with schizophrenia recruited in Hanoi, Vietnam. Moreover, we aim to facilitate genetic discoveries by contributing to global research endeavors to diversify genetic samples and clinical data in schizophrenia research. Ultimately, this pilot study sets the foundation for the recruitment of a sizeable Vietnamese well characterized cohort that includes state-of-the-art phenotyping procedures in the field of psychiatric genetics.

## 2 Methods

### 2.1 Local Recruitment and Data Collection

For this pilot study, 190 patients with schizophrenia and 188 control subjects, all of Vietnamese Kinh ancestry, were recruited between August and December 2017. All participants were at least 18 years old and provided full written informed consent upon study enrollment. Before data collection, study procedures were reviewed and approved by the institutional ethical review board of the Hanoi Medical University. All in-patients were recruited at teaching hospitals of the Department of Psychiatry at Hanoi Medical University.

Additionally, outpatients were recruited from associated community medical centers around the Hanoi metropolitan area. In outpatient facilities, data collection was supervised by senior psychiatrists of the staff at the Department of Psychiatry, Hanoi Medical University. Diagnosis of schizophrenia according to ICD-10 criteria (F20.X) was ascertained by clinicians with at least three years of psychiatric experience and confirmed by a senior attending psychiatrist. Current symptom severity was assessed with the Vietnamese Translation of the Positive and Negative Syndrome Scale (Kay et al., 1987), which encompasses the dimensions of positive, negative, and general psychopathology. Each patient was interviewed and rated independently by two clinicians who received prior training in conducting the semi-structured interview. If no consensus was reached, ratings were discussed with a senior psychiatrist. Medical and demographic parameters, including medication, duration of illness, and age at onset, were recorded during routine examinations and/or extracted from the patients’ medical chart. Control subjects were recruited from the staff at the Hanoi Medical University across different occupational groups. To obtain genetic information, saliva samples were collected from each subject using OraGene-510 DNA-Self-Collection Kits (Genotek, Ottawa, Ontario, Canada) according to the manufacturer’s instructions.

### 2.2 Genetic Data

#### 2.2.1 Genome-wide Genotyping

Genetic material was extracted from saliva following established standard protocols. Genome-wide genotyping of DNA samples was performed on Infinitum Global Screening array-23 (GSA) MD BeadChip (Illumina, San Diego, CA) at ERASMUS Medical Center, Human Genotyping Facility (HuGe-F), Rotterdam, Netherlands. SNP-level data was analyzed using a central pipeline ‘RICOPILI’ (Lam et al., 2020) on the LISA computer cluster.

#### 2.2.2 Quality Control

According to the Psychiatric Genomics Consortium (PGC) standards, quality control was performed on the newly recruited cohort. The default quality control parameters for retaining subjects and SNPs were: SNP missingness < 0.05 (before sample removal); subject missingness < 0.02; autosomal heterozygosity deviation (| F_het_ | < 0.2); SNP missingness < 0.02 (after sample removal); difference in SNP missingness between cases and controls < 0.02; and SNP Hardy-Weinberg equilibrium (*P* > 10^−6^ in controls or *P* > 10^−10^ in cases). Seven individuals who did not meet QC criteria were excluded resulting in a data set of 370 individuals (184 cases and 186 controls) and 367,920 SNPs. Post-QC Manhattan and QQ plots did not show unexpected genome-wide significant association or general inflation.

#### 2.2.3 Genotype Imputation

Genotype imputation was performed using the pre-phasing/imputation stepwise approach implemented in EAGLE/MINIMAC3 (with a variable chunk size of 132 genomic chunks and default parameters). The imputation reference set consisted of 54,330 phased haplotypes with 36,678,882 variants from the publicly available HRC reference release 1.1 (McCarthy et al., 2016).

#### 2.2.4 Population Stratification and Relatedness Testing

After imputation, we identified SNPs with very high imputation quality (INFO >0.8) and low missingness (<1%) for further quality control. After linkage disequilibrium pruning (r^2^ > 0.02) and frequency filtering (MAF > 0.05), there were 116,203 autosomal SNPs in the data set. This SNP set was used for robust relatedness testing and population structure analysis using PLINK (Purcell et al., 2007). Thirteen pairs of subjects with identity-by-descent PI-HAT value of > 0.2 were identified. One member of each pair was removed at random after preferentially retaining cases over controls. Principal component estimation was done with the same collection of autosomal SNPs, and fifteen ancestry outliers were removed from the data set (see supplementary figures S2-S3). Supplementary figure S3 (A) shows that Vietnamese individuals cluster within other Asian populations from the 1000 Genomes reference population (Auton et al., 2015). Figure S3 (B) illustrates that the Vietnamese Kinh population is distinctly stratified from China and Japan when plotted only against Asian populations. We tested the first 20 principal components for phenotype association (using logistic regression with study indicator variables included as covariates). Seven principal components, namely PC1, PC2, PC3, PC4, PC5, PC8, and PC14, were significantly associated (P<0.05) with SCZ status, and these were added to the GWAS model as covariates in order to control for population stratification. The final analysis consisted of 343 individuals (174 cases and 169 controls) and 8,332,179 SNPs. We tested all SNPs for association under an additive logistic regression model using PLINK and the derived principal components as covariates.

#### 2.2.5 Polygenic Risk Scoring

We obtained summary statistics from the latest PGC schizophrenia GWAS (Schizophrenia Working Group of the Psychiatric Genomics Consortium et al., 2020) to calculate PRS for individuals in our target data set. Polygenic scoring was performed using (a) East-Asian (EAS) specific meta-analysis (14,004 cases and 16,757 controls), (b) European (EUR) specific meta-analysis (53,386 cases and 77,258 controls) and (c) trans-ancestry (MIX) meta-analysis (67,390 cases and 94,015 controls) as training statistics. Genome-wide summary statistics were LD pruned and “clumped” by discarding variants within 500 kb of, and in r2 ≥ 0.1 with another (more significant) marker, resulting in 61 805 (EAS), 90,713, (EUR) 87,671 variants (MIX), respectively. Clumping for the East-Asian ancestry was conducted with the LD information of the East-Asian subset of the HRC reference panel. The EUR and MIX training data were clumped using LD information from the entire HRC reference panel. This set of LD-independent SNPs was used to construct a PRS at a range of P-value thresholds (5×10^−8^, 1×10^−6^, 1×10^−4^, 0.001, 0.01, 0.05, 0.1, 0.2, 0.5, 1.0). For a given sample, we take each LD-independent SNP and multiply the marginal SNP effect from the logistic regression by the number of risk alleles. The resulting products were summed over all variants for each individual so that a single score reflecting the overall polygenic burden for schizophrenia is obtained.

#### 2.2.6 Consistency of Directions of Effects

In an additional final step, we assessed the robustness of genetic associations in the Vietnamese pilot sample by performing a replication analysis using the PGC trans-ancestry discovery sample as a discovery data set. We used a binomial sign test to establish that SNP associations replicated *en masse* in an independent PGC sample at various p-value thresholds by showing that the number of concordant effect directions for index SNPs was significantly greater than the null hypothesis of randomly oriented effects. The Vietnam summary statistics were then combined with those from the PGC primary GWAS dataset using an inverse variance-weighted fixed effects model and overlapping LD-clumps were merged into distinct genomic regions.

### 2.3 Polygenic Association Testing

Individual-level PRS at each p-value threshold (Pd) were tested for association with SCZ status (Schizophrenia vs. No Schizophrenia) in logistic regression models. To adjust for fine-grained, population-stratification principal components (PCs) 1, 2, 3, 4, 5, 8, and 14 (selected for being significantly associated with SCZ status) were included as covariates in each model. Nagelkerke’s R^2^, as a measure of “variance explained” for logistic regression, was determined by comparing the full model (with PRS and PCs) against the null model containing only PCs. We report Nagelkerke’s R^2^ on the liability scale (Lee et al., 2012), which corrects for a higher case-control-ratio in the target dataset relative to the general population’s prevalence estimated 0.7% (McGrath et al., 2008). Using LD-clumped genome-wide summary statistics without any p-value filter, individuals were grouped into five quintiles based on their individual polygenic burden (SCZ-PRS). Odds ratios for schizophrenia diagnosis were obtained by comparing individuals with low polygenic risk (bottom 20%, or bottom quintile) to individuals in strata with increased polygenic loadings for SCZ (second to the fifth quintile). Each step was repeated for three different training data sets: East-Asian GWAS (EAS), European GWAS (EUR), and mixed-ancestry GWAS (PGC3 SCZ-GWAS).

## 3 Results

### 3.1 Sample Characteristics

A total of 378 participants were included in the study, of which 190 are cases diagnosed with schizophrenia and 188 are control subjects. Sociodemographic parameters of all case and control subjects are shown in Table 1. All clinical parameters of the case sample are presented in Table 2.

**Table 1.** Sociodemographic characteristics of the health control and patient sample

|  |  | **Vietnamese sample (N=378)** |  |
| --- | --- | --- | --- |
|  |  | **Patients (N=190)** | **Controls (N=188)** |
| **Age** [years] | Mean (SD) | 44.44 (10.62) | 44.21 (10.43) |
| **Sex** (%) |  |  |  |
|  | Male | 147 (77.37) | 134 (71.28) |
|  | Female | 43 (22.63) | 54 (28.72) |
| **Weight** [kg] | Mean (SD) | 57.92 (9.81) | 62.46 (8.97) |
| **Height** [cm] | Mean (SD) | 162.45 (7.35) | 164.34 (7.14) |
| **BMI** [kg/m<sup>2</sup>] | Mean (SD) | 21.90 (3.33) | 23.06 (2.40) |
| **Marital Status** (%) |  |  |  |
|  | Single | 97 (51.32) | 9 (4.86) |
|  | Married | 58 (30.69) | 172 (92.97) |
|  | Living separately | 10 (5.29) | 1 (0.54) |
|  | Divorced | 21 (11.11) | 0 (0.00) |
|  | Widowed | 3 (1.59) | 3 (1.62) |
| **School Education** (%) |  |  |  |
|  | Illiterate | 2 (1.06) | 0 (0.00) |
|  | Primary School | 29 (15.4) | 1 (0.53) |
|  | Secondary School | 59 (31.38) | 21 (11.23) |
|  | High School | 100 (53.19) | 165 (88.24) |
| **Higher Education** (%) |  |  |  |
|  | Apprentice | 12 (27.91) | 6 (4.32) |
|  | Under-College | 9 (20.93) | 40 (28.78) |
|  | College / University | 22 (51.65) | 67 (48.22) |
|  | Post-University | 0 (0.00) | 26 (18.71) |

**Table 2.** Clinical parameters

|  |  | N (%) | Mean (SD) |
| --- | --- | --- | --- |
| **Clinical information** |  |  |  |
| Duration of illness [years] |  |  | 18.72 (10.09) |
| Average age of onset of positive symptoms |  |  | 25.55 (7.52) |
| No. of hospitalisations<sup>1</sup> |  |  | 11.51 (15.50) |
| Positive Family History<sup>2</sup> | at least 1 family member affected | 54 (28.42) |  |
| Recruitment site | Inpatients | 96 (50.53) |  |
|  | Outpatients | 94 (49.47) |  |
| **Treatment** |  |  |  |
| Antipsychotic Medication<sup>3</sup> | Olanzapine | 59 (31.05) |  |
|  | Risperidone | 14 (7.37) |  |
|  | Clozapine | 12 (6.32) |  |
|  | Haloperidol | 7 (3.68) |  |
|  | Chlorpromazine | 11 (5.79) |  |
|  | Levomepromazine | 68 (35.79) |  |
|  | Other antipsychotics | 17 (9.45) |  |
| **Symptom severity** |  |  |  |
| Symptoms within the last year | Complete remission | 1 (0.53) |  |
|  | Currently symptomatic | 188 (98.95) |  |
| PANSS | PANSS-Pos |  | 14.80 (6.50) |
|  | PANSS-Neg |  | 24.42 (8.81) |
|  | PANSS-Gen |  | 34.01 (12.08) |
|  | PANSS-Total |  | 73.12 (22.52) |
| Domains of psychopathology | Delusions | 75 (39.47) |  |
|  | Hallucinations | 80 (42.11) |  |
|  | Disorganisation | 77 (40.53) |  |
|  | Catatonia | 21 (11.05) |  |
|  | Negative symptoms | 189 (99.47) |  |
|  | Hostile behavior | 58 (30.53) |  |
|  | Marked affective symptoms | 21 (11.05) |  |
|  | Other psychopathological symptoms | 5 (2.63) |  |

<sup>1</sup> Number of lifetime in-patient admissions due to schizophrenia<sup>2</sup> Family history of psychiatric illnesses<sup>3</sup> All patients received at least one antipsychotic medication

### 3.2 Polygenic Risk Profiling

Next, we assessed how much variance in schizophrenia risk could be attributed to underlying genetic variation. After quality control (see **Supplementary** material), 343 individuals remained for polygenic risk score analysis. Schizophrenia polygenic risk scores (SCZ-PRS) were generated with GWAS summary statistics from East-Asian (EAS), European (EUR), and mixed-ancestry populations as training sets. In line with previous findings, individual-level SCZ-PRS were significantly associated with schizophrenia case status (**Fig 1, Supplementary Table S1**). However, the predictive accuracy of SCZ-PRS among Vietnamese participants varied with the ancestral composition of the training data sets. The strongest effect sizes were observed when PRS were derived from trans-ancestry summary statistics, which explained ∼4.9 % of the variance in schizophrenia liability (p = 6.83 x 10^−8^ at Pd < 1). Likewise, PRS constructed on East-Asian GWAS summary statistics (PGC SCZ wave 3) accounted for ∼4.5 % of the variability in schizophrenia risk (p = 2.73 x 10^−7^ at Pd < 1). PRS trained on European ancestry GWAS results also predicted SCZ status in the Vietnamese target sample with an explained variance of only ∼3.9 % (p = 1.79 x 10^−6^ at Pd < 0.01). As expected, best overall predictions were observed at more inclusive p-value thresholds that capture a larger proportion of the polygenic signal by including more independent common variants below the genome-wide significance level.

**Fig 1.**
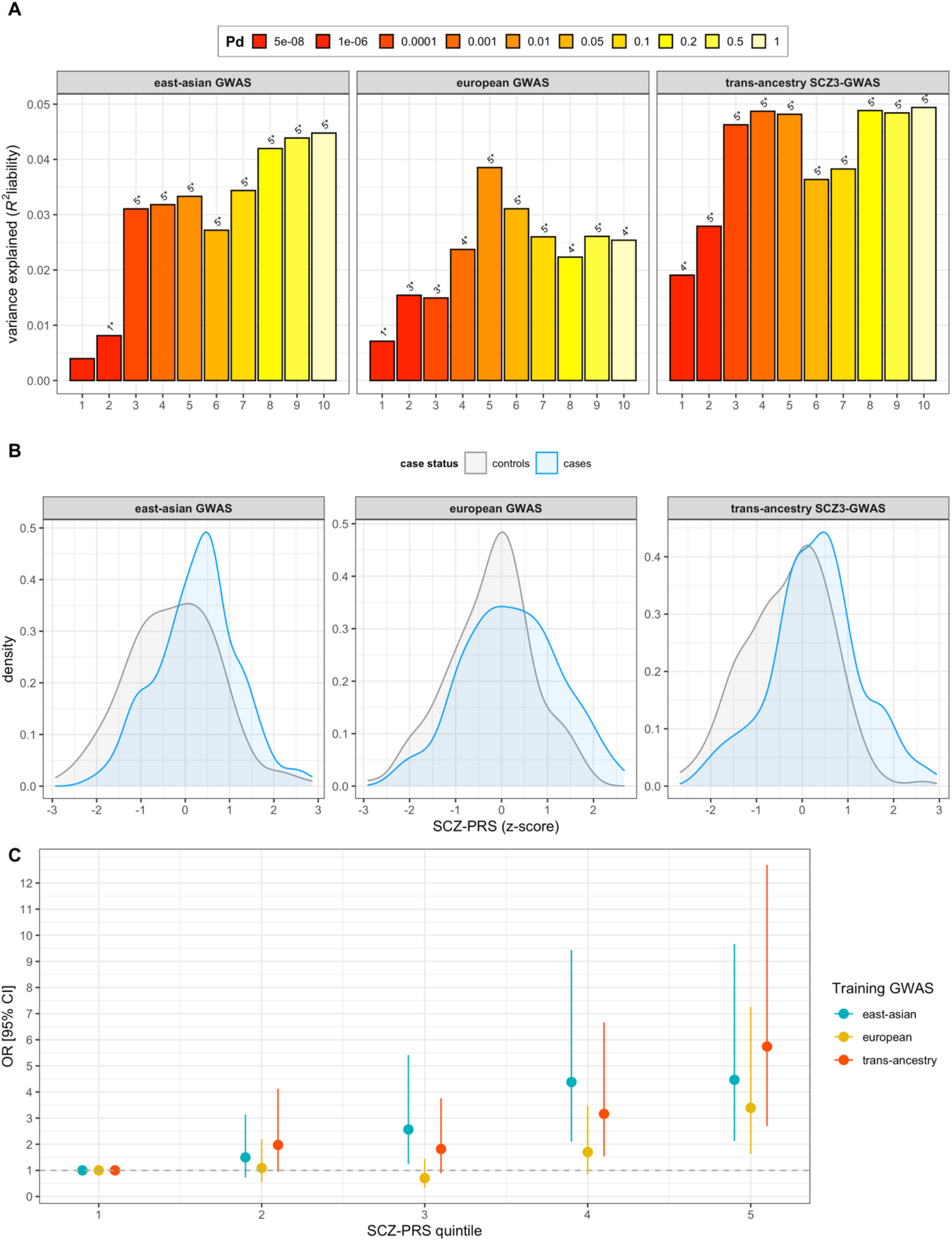
Associations of SCZ-PRS with schizophrenia in a cohort of 169 controls and 174 cases of Vietnamese Kinh ancestry. **A)** The variance explained (R^2^ liability) by SCZ-PRS at different p-value thresholds (Pd; x-axis) is shown on the y-axis. P-values are displayed on top of each bar: 1* < 0.05, 2* < 0.01, 3* < 0.005, 4* <0.001, 5* < 1×10^−4^. SCZ-PRS were constructed on different GWAS discovery samples: East-Asian GWAS (left panel), European-ancestry GWAS (center panel), and trans-ancestry SCZ3-GWAS (right panel). **B)** Distribution of z-transformed polygenic risk scores among cases and controls at Pd < 1. **C)** odds ratios for schizophrenia per SCZ-PRS (Pd < 1) quintile relative to the first quintile. Error bars represent 95% Confidence Intervals.

A similar pattern can be observed when inspecting odds ratios for the association between SCZ-PRS and schizophrenia diagnosis (**Fig 1c, Supplementary Table S3**). Overall, individuals who carry a higher polygenic burden are at higher odds for having a schizophrenia diagnosis, ORs range from 1.97 (95% CI, 0.96 - 4.12) at the second PRS-quintile up to 5.74 (95% CI, 2.70 - 12.69) at the fifth PRS-quintile, using the first quintile as the reference group and mixed-ancestry training populations for polygenic scoring. When considering scores obtained from east-Asian ancestry GWAS, odds ratios for SCZ are slightly lower, ranging from 1.50 (95% CI, 0.72 - 3.14) at the second PRS-quintile up to 4.47 (95% CI, 2.13 – 9.66) at the top PRS-quintile. By comparison, observed ORs are markedly lower for EUR-derived SCZ-PRS; the OR at the fifth PRS quintile is 3.39 (95% CI, 1.63 – 7.26) relative to the first PRS quintile.

Overall, reported predictive values of PRS and odds ratios at the top PRS-quintile in our Vietnamese sample are consistent with those of other East-Asian cohorts reported within the latest SCZ GWAS meta-analysis (Schizophrenia Working Group of the Psychiatric Genomics Consortium, 2020).

### 3.3 Replication Analysis

Complementing PRS association testing, we explored if SNP associations at varying p-value thresholds (Pd) in the mixed-ancestry SCZ3-GWAS replicate in the present Vietnamese-ancestry sample (replication data set). We observed that the fraction of SNPs with directions of effect congruent to the replication data set was consistently greater than 50%. Specifically, of the 182 index SNPs that attained genome-wide significance in the SCZ3-GWAS meta-analysis and that were also available in the Vietnamese cohort, 110 SNPs (60%) showed the same direction of effect, which was significantly greater than expected by chance (binomial p-value = 0.003). Results of the replication analysis and binomial sign test across different p-value thresholds are reported in the supplementary material (**Supplementary Table S3)**. When meta-analyzed with SCZ3-GWAS, 249 loci reach genome-wide significance, in contrast to 244 loci identified by SCZ3-GWAS alone, indicating that by adding this or a potentially larger Vietnamese cohort could help to discover novel SCZ-associated loci in future GWAS meta-analysis.

## 4 Discussion

With this pilot study, the feasibility of a large-scale data collection of patients diagnosed with schizophrenia in Vietnam has been shown for the first time. We were able to implement a successful recruitment scheme at the Department of Psychiatry of the Hanoi Medical University, with the National Institute of Mental Health staff at Bach Mai Hospital and the Hanoi Psychiatric Hospital participating in data collection. This study established an efficient pipeline for data generation and subsequent analyses that will be upscaled with future collaborative endeavors. With these central foundations set in place, we aim to continue and intensify our data collection efforts and establish more in-depth phenotyping going forward.

There are a few practical and clinical considerations concerning the recruitment of patients with schizophrenia in Vietnam. First, patients were primarily recruited through in-patient facilities. For genetic research, that may prove to be an advantage since the clinical population in Vietnam comprises mostly cases with severe illness courses of schizophrenia, long duration of illness, multiple prior hospitalizations, and a high symptom load, especially for negative and residual symptoms (Maramis et al., 2011). This pattern can be considered representative of inpatients often treated for long durations for chronic symptoms of schizophrenia in Vietnam, since individuals with a less severe course of illness typically won’t seek or receive hospital treatment due to stigma or a lack of illness recognition for less severe illness courses (Martensen et al., 2020; Nguyen et al., 2019; Ta et al., 2016, 2018). This pattern is reflected by the fact that even today, more than 50% of all patients treated in psychiatric hospitals in Vietnam have a primary psychiatric diagnosis of schizophrenia (Maramis et al., 2011; Martensen et al., 2018, 2020). In our pilot sample, most of the patients were hospitalised several times due to schizophrenia. Nearly all patients, including the outpatients, still exhibited chronic, residual, or negative symptoms when assessed by the PANSS at the time of recruitment. To increase clinical heterogeneity among patients, we will integrate more outpatient facilities in the Hanoi metropolitan area as additional study sites in future studies.

In the PRS and replication analysis, we corroborated some key findings of previous publications. First, we observed that PRS derived from trans-ancestry GWAS perform better than EAS-derived PRS, and both predict schizophrenia more accurately than EUR-derived PRS. Similarly, leave-one-out PRS analysis in the latest SCZ GWAS meta-analysis demonstrated that PRS in Asian cohorts explained consistently more variance in schizophrenia risk when mixed-ancestry summary statistics were used for polygenic scoring compared to Asian training populations (Schizophrenia Working Group of the Psychiatric Genomics Consortium et al., 2020). Consistent with this finding, Lam et al. (2019) demonstrated that EUR-derived PRS accounted for less variance in schizophrenia liability than EAS-derived PRS. Second, we demonstrated that risk-conferring SNPs in the present Vietnamese samples largely showed same-direction effects compared to SCZ PGC3 summary statistics. The inclusion of our sample, especially after scaling-up the sample size, will probably uncover novel susceptibility loci, especially since an earlier meta-analysis of East-Asian (Lam et al., 2019) and Latino and admixed African samples (Bigdeli et al., 2020) strongly support this notion. Due to practical limitations, we focused on Kinh ancestry individuals, representing 87% of the Vietnamese population. Given the genetic diversity among Vietnamese populations (Macholdt et al., 2020), our genotyping initiative might be extended to other ethnic groups living in Vietnam and bordering geographical areas in South-East-Asia. Thus, the addition of currently understudied populations such as Vietnamese ethnicities will likely help refine our understanding of the genetic architecture of schizophrenia.

Ancestral diversity among study participants in discovery GWAS has a substantial impact on prediction accuracy of polygenic scores and their applicability across populations. Further inclusion of East- and South-East-Asian cohorts will likely improve the polygenic risk prediction in Vietnamese individuals and vice versa, more than by a comparable addition of European ancestry samples. This is increasingly important as PRS are beginning to find their way into clinical practice outside of psychiatry. Beyond disease risk prediction, the clinical utility of PRS may range from genotype-informed screening methods and stratification to tailored intervention strategies (Torkamani et al., 2018). To translate genome-wide data into clinically actionable information, in-depth clinical and valid phenotypic information must be collected along with genetic data. While these pilot data are too small to perform meaningful secondary analyses, e.g., on symptom profiles, such cohorts will play a pivotal role in making potential PRS-based applications available for a wider population.

## 5 Conclusion and Outlook

This pilot research project’s main objective was to establish key research and clinical infrastructures for a larger cross-sectional Vietnamese cohort of patients with schizophrenia and healthy controls. In the next phase, we will extend our data collection efforts by introducing additional study sites in Vietnam, both in northern and southern provinces, as well as a translated and validated comprehensive phenotyping battery. This includes trained assessment, that is harmonised with our current research initiative in Berlin, Germany. This assessment is comprised of an in-depth neuropsychological evaluation and a selection of questionnaires that assess environmental risk factors, a broad range of health-related data, as well as outcome measures such as quality of life and social functioning. With this prospective cohort, we intend to build a valuable resource for trans-ancestry GWAS and secondary analyses on cross-cultural gene-environment interactions to elucidate potential etiological trajectories associated with an increased liability to Schizophrenia. In doing so, we aim to contribute to a worldwide endeavor to enhance well-powered and ancestrally diverse GWAS samples and explore trans-ancestry and culture specific gene-environment interactions in psychiatry.

## Data Availability

Individual-level genotypes are available from the corresponding author upon reasonable request.

## Acknowledgements

We thank the Schizophrenia Working Group of the Psychiatric Genomics Consortium for granting permission to use European, East-Asian and trans-ancestry GWAS summary statistics (wave 3) for this study. The Stanley Center for Psychiatric Research financially supported the collection of this sample. The statistical analysis was carried out on the Dutch LISA computer cluster powered by SURFsara. We further thank the participants and additional staff of Hanoi Psychiatric Hospital, Hanoi Medical University, and the National Institute of Mental Health, Vietnam, for taking part in this study.

