## Supplemental material for "Increasing Sample Diversity in Psychiatric Genetics – Introducing a new Cohort of Patients with Schizophrenia and Controls from Vietnam – Results from a Pilot Study"

### SUPPLEMENTARY MATERIAL

*Technical Quality Control*

A. B.

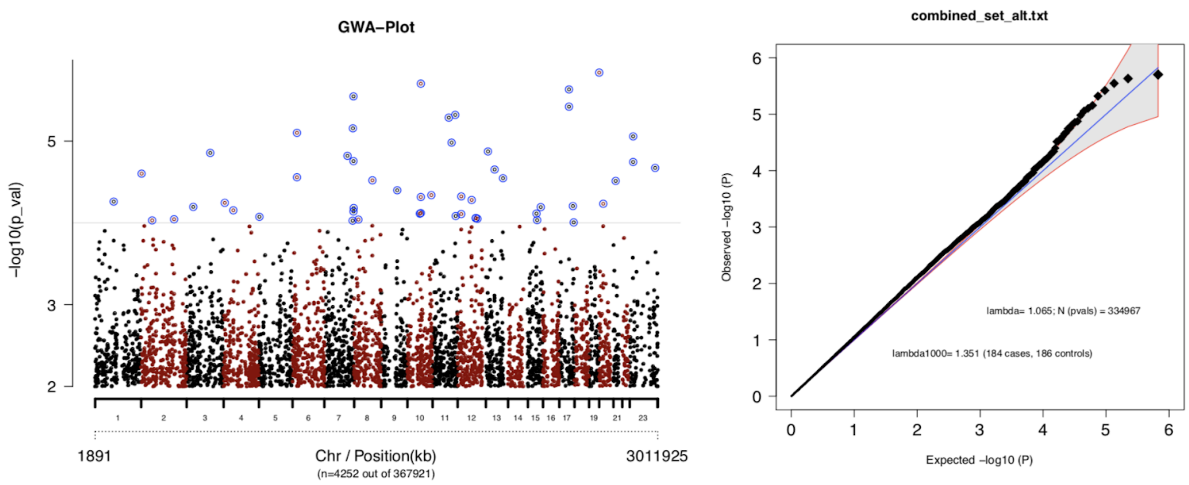


**Supplementary Figure S1.** A. Manhattan plot after initial quality control. Chromosomes position in ascending order is indicated on the Y axis, the -log$_{10}$(p-values) of included autosomal SNPs derived from the association analysis are plotted on the y axis. B. Quantile-quantile (Q-Q) plot of observed plotted against expected –log$_{10}$(P-values) sorted from smallest to largest –log$_{10}$(P-values) and Lambda statistic before excluding population outliers.

*Genomic Quality Control*

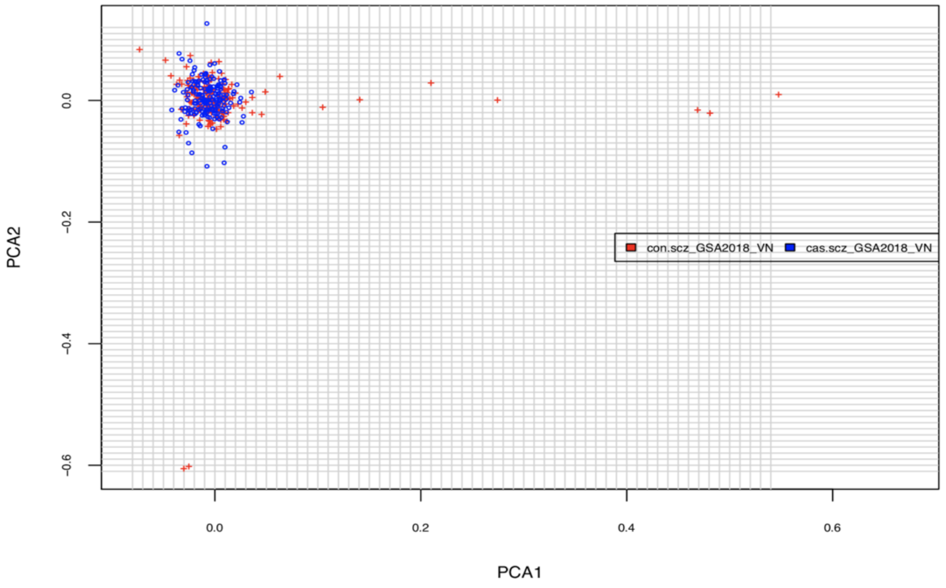


**Supplementary Figure S2**. Principal Component Analysis (PCA) Plot of the first two principal components reflecting the presence of population structure in the sample. Individuals with PC1 > 0.1 and -0.06 < PC2 > 0.1 were considered to be ancestry outliers and have been excluded from further analyses.

A.

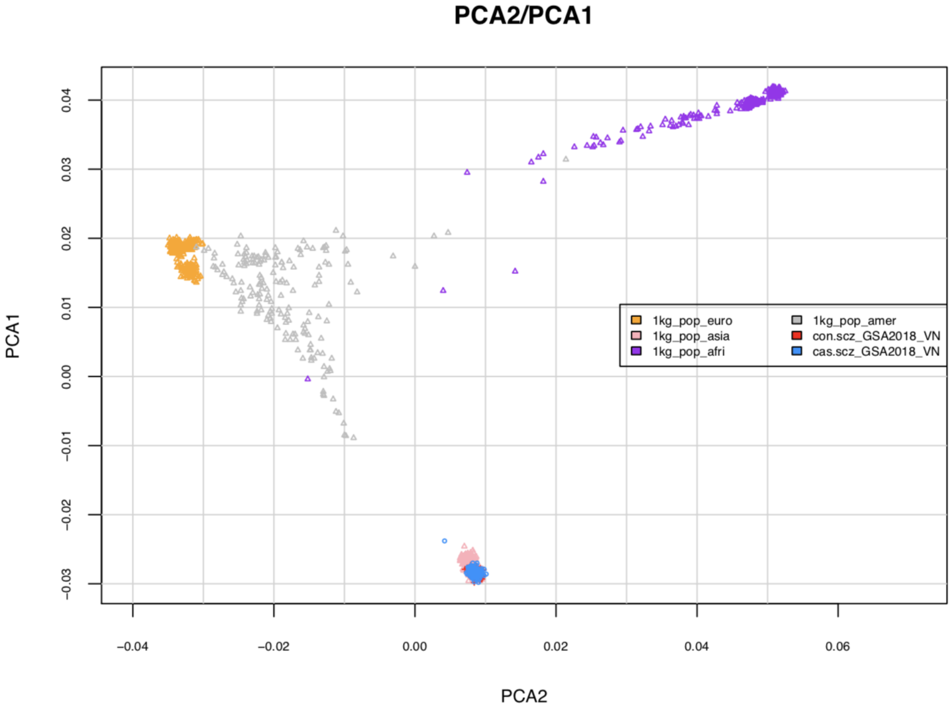


B.

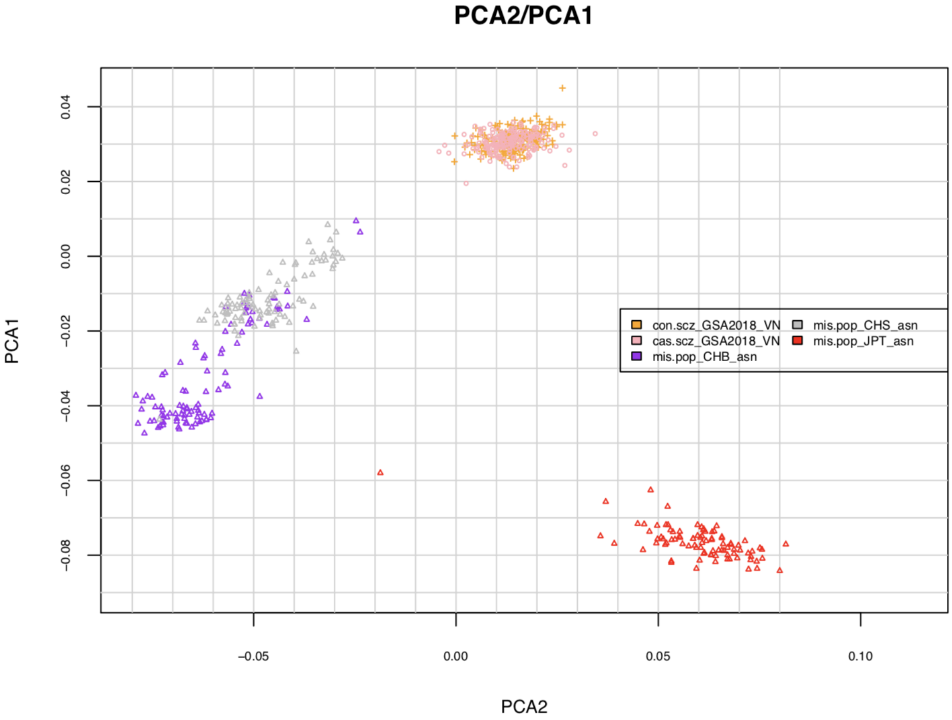


**Supplementary Figure S3**. PCA-clustering of Vietnamese individuals anchored by 1000 Genomes reference populations. A) Vietnamese samples are mapped to Asian, African, American and European samples. Vietnamese cases are colored in blue, controls in red. B) Vietnamese samples are mapped to Japanese (JPT), Han Chinese (CHB) and Southern Han Chinese (CHS) populations. Cases are colored in pink, controls in yellow.

*Polygenic risk scoring*

**Supplementary Table S1**. Associations of SCZ-PRS with schizophrenia in the Vietnamese target sample. For each training data set (EAS, EUR, MIX), the number of SNPs on which PRS are constructed on, variance explained (Nagelkerke's $R^2$, $R^2$ liability scale), area under the curve (AUC) and p-values are reported at each p-value threshold (Pd)

| **Pd** | | **EAS** (16,757 controls, 14,004 cases) | **EUR** (77,258 controls, 53,386 cases) | **MIX** (94,015 controls, 67,390 cases) |
|---|---|---|---|---|
| 0.00000005 | **SNPs** | 9 | 206 | 268 |
| | **Observed Nagelkerke $R^2$** | 0.0095 | 0.0170 | 0.0449 |
| | **p-value** | 0.1232 | 0.0388 | 0.00074 |
| | **$R^2$ liability** | 0.0039 | 0.0071 | 0.0191 |
| | **$R^2$ liability (SE)** | 0.0050 | 0.0064 | 0.0106 |
| | **AUC** | 0.556 | 0.574 | 0.609 |
| 0.000001 | **SNPs** | 23 | 411 | 515 |
| | **Observed Nagelkerke $R^2$** | 0.0194 | 0.0365 | 0.0650 |
| | **p-value** | $2.73 \times 10^{-2}$ | $2.38 \times 10^{-3}$ | $04.61 \times 10^{-5}$ |
| | **$R^2$ liability** | 0.0081 | 0.0154 | 0.0280 |
| | **$R^2$ liability (SE)** | 0.0072 | 0.0094 | 0.0126 |
| | **AUC** | 0.580 | 0.604 | 0.627 |
| 0.0001 | **SNPs** | 198 | 1620 | 1891 |
| | **Observed Nagelkerke $R^2$** | 0.0721 | 0.0354 | 0.1053 |
| | **p-value** | $1.75 \times 10^{-5}$ | $2.80 \times 10^{-3}$ | $1.74 \times 10^{-7}$ |
| | **$R^2$ liability** | 0.0311 | 0.0149 | 0.0463 |
| | **$R^2$ liability (SE)** | 0.0134 | 0.0091 | 0.0159 |
| | **AUC** | 0.619 | 0.598 | 0.669 |
| 0.001 | **SNPs** | 846 | 4000 | 4303 |
| | **Observed Nagelkerke $R^2$** | 0.0734 | 0.0555 | 0.1106 |
| | **p-value** | $1.39 \times 10^{-5}$ | $1.71 \times 10^{-4}$ | $8.38 \times 10^{-8}$ |
| | **$R^2$ liability** | 0.0318 | 0.0237 | 0.0487 |
| | **$R^2$ liability (SE)** | 0.0137 | 0.0114 | 0.0164 |
| | **AUC** | 0.627 | 0.615 | 0.671 |
| 0.01 | **SNPs** | 4170 | 11000 | 11500 |
| | **Observed Nagelkerke $R^2$** | 0.0771 | 0.0886 | 0.1094 |
| | **p-value** | $8.74 \times 10^{-6}$ | $1.79 \times 10^{-6}$ | $9.85 \times 10^{-8}$ |
| | **$R^2$ liability** | 0.0333 | 0.0385 | 0.0482 |

| | | | | |
|---|---|---|---|---|
| | **$R^2$ liability (SE)** | 0.0140 | 0.0147 | 0.0164 |
| | **AUC** | 0.644 | 0.643 | 0.656 |
| 0.05 | **SNPs** | 12100 | 24100 | 24700 |
| | **Observed Nagelkerke $R^2$** | 0.0634 | 0.0721 | 0.0838 |
| | **p-value** | $5.79 \times 10^{-5}$ | $1.74 \times 10^{-5}$ | $3.45 \times 10^{-6}$ |
| | **$R^2$ liability** | 0.0272 | 0.0311 | 0.0364 |
| | **$R^2$ liability (SE)** | 0.0126 | 0.0134 | 0.0144 |
| | **AUC** | 0.624 | 0.637 | 0.649 |
| 0.1 | **SNPs** | 18900 | 34400 | 34700 |
| | **Observed Nagelkerke $R^2$** | 0.0794 | 0.0607 | 0.0880 |
| | **p-value** | $6.34 \times 10^{-6}$ | $8.40 \times 10^{-5}$ | $1.93 \times 10^{-6}$ |
| | **$R^2$ liability** | 0.034 | 0.0260 | 0.0382 |
| | **$R^2$ liability (SE)** | 0.0140 | 0.0124 | 0.0149 |
| | **AUC** | 0.642 | 0.622 | 0.649 |
| 0.2 | **SNPs** | 29200 | 48900 | 48200 |
| | **Observed Nagelkerke $R^2$** | 0.0961 | 0.0523 | 0.111 |
| | **p-value** | $6.32 \times 10^{-7}$ | $2.64 \times 10^{-4}$ | $8.07 \times 10^{-8}$ |
| | **$R^2$ liability** | 0.0420 | 0.0223 | 0.489 |
| | **$R^2$ liability (SE)** | 0.0154 | 0.0115 | 0.0168 |
| | **AUC** | 0.654 | 0.606 | 0.664 |
| 0.5 | **SNPs** | 48700 | 73800 | 72100 |
| | **Observed Nagelkerke $R^2$** | 0.100 | 0.0609 | 0.12 |
| | **p-value** | $3.57 \times 10^{-7}$ | $8.17 \times 10^{-5}$ | $9.18 \times 10^{-8}$ |
| | **$R^2$ liability** | 0.0439 | 0.0261 | 0.0484 |
| | **$R^2$ liability (SE)** | 0.0156 | 0.0124 | 0.0166 |
| | **AUC** | 0.657 | 0.620 | 0.667 |
| 1 | **SNPs** | 61800 | 90700 | 87700 |
| | **Observed Nagelkerke $R^2$** | 0.1021 | 0.0507 | 0.112 |
| | **p-value** | $2.73 \times 10^{-7}$ | $1.01 \times 10^{-4}$ | $6.83 \times 10^{-8}$ |
| | **$R^2$ liability** | 0.0448 | 0.0253 | 0.049 |
| | **$R^2$ liability (SE)** | 0.0156 | 0.0122 | 0.0168 |
| | **AUC** | 0.654 | 0.618 | 0.667 |

**Supplementary table S2**. Odds ratios (ORs) for schizophrenia at second to fifth SCZ-PRS quintile. The first SCZ-PRS quintile serves as a reference group. Number of controls ($N_{con}$) and number of cases ($N_{cas}$) are reported for each quintile.

| Training data set | Quintile | $N_{con}$ | $N_{cas}$ | OR | 95% CI | p-value |
|---|---|---|---|---|---|---|
| European GWAS | 1 | 39 | 30 | 1.00 | (1.00 - 1.00) | - |
| | 2 | 37 | 32 | 1.09 | (0.54 - 2.20) | $8.10 \times 10^{-1}$ |
| | 3 | 45 | 24 | 0.70 | (0.34 - 1.44) | $3.37 \times 10^{-1}$ |
| | 4 | 29 | 39 | 1.70 | (0.84 - 3.48) | $1.42 \times 10^{-1}$ |
| | 5 | 19 | 49 | 3.39 | (1.63 - 7.26) | $1.31 \times 10^{-3}$ |
| Trans-ancestry SCZ3-GWAS | 1 | 47 | 22 | 1.00 | (1.00 - 1.00) | - |
| | 2 | 37 | 32 | 1.97 | (0.96 - 4.12) | $6.79 \times 10^{-2}$ |
| | 3 | 38 | 31 | 1.82 | (0.89 - 3.76) | $1.04 \times 10^{-1}$ |
| | 4 | 28 | 40 | 3.16 | (1.54 - 6.67) | $2.04 \times 10^{-3}$ |
| | 5 | 19 | 49 | 5.74 | (2.70 -12.69) | $9.12 \times 10^{-6}$ |
| East-Asian GWAS | 1 | 47 | 22 | 1.00 | (1.00 - 1.00) | - |
| | 2 | 41 | 28 | 1.50 | (0.72 - 3.14) | $2.79 \times 10^{-1}$ |
| | 3 | 33 | 36 | 2.56 | (1.24 - 5.42) | $1.20 \times 10^{-2}$ |
| | 4 | 25 | 43 | 4.38 | (2.10 - 9.44) | $1.13 \times 10^{-4}$ |
| | 5 | 23 | 45 | 4.47 | (2.13 - 9.66) | $1.01 \times 10^{-4}$ |

*Replication Analysis*

**Supplementary table S3.** Binomial sign test for consistency of directions of allelic effect between Vietnamese cohort and SCZ3-GWAS

| Replication data set | Pd | No of rep. SNPs | No of SNPs | Sign test p-value | Ratio |
|---|---|---|---|---|---|
| | $1 \times 10^{-4}$ | 1198 | 2197 | $1.18 \times 10^{-5}$ | 0.55 |
| Trans-ancestry SCZ3-GWAS | $1 \times 10^{-5}$ | 577 | 1021 | $1.76 \times 10^{-5}$ | 0.57 |
| | $1 \times 10^{-6}$ | 315 | 529 | $6.50 \times 10^{-6}$ | 0.6 |
| | $5 \times 10^{-7}$ | 170 | 275 | $5.31 \times 10^{-5}$ | 0.62 |
| | $1 \times 10^{-7}$ | 116 | 192 | $2.38 \times 10^{-3}$ | 0.6 |
| | $5 \times 10^{-8}$ | 110 | 182 | $2.97 \times 10^{-3}$ | 0.6 |
